# Age Prediction From 12-lead Electrocardiograms Using Deep Learning: A Comparison of Four Models on a Contemporary, Freely Available Dataset

**DOI:** 10.1101/2024.02.02.24302201

**Authors:** Andrew Barros, Ian German-Mesner, N. Rich Nguyen, J. Randall Moorman

## Abstract

**Objective:** The 12-lead electrocardiogram (ECG) is routine in clinical use and deep learning approaches have been shown to have the identify features not immediately apparent to human interpreters including age and sex. Several models have been published but no direct comparisons exist.

**Approach:** We implemented three previously published models and one unpublished model to predict age and sex from a 12-lead ECG and then compared their performance on an open-access data set.

**Main results:** All models converged and were evaluated on the holdout set. The best preforming age prediction model had a hold-out set mean absolute error of 8.06 years. The best preforming sex prediction model had a hold-out set area under the receiver operating curve of 0.92.

**Significance:** We compared performance of four models on an open-access dataset.

## INTRODUCTION

The electrocardiogram (ECG) entered clinical use in the early 1900’s and earned Willem Einthoven a Nobel prize in 1924(Pahlm and Uvelius 2019). For much of the last 100 years, the ECG has been acquired by a machine, printed in a canonical paper form, and interpreted by physicians. Many conditions manifest as alterations of the ECG. In the 1980’s automated interpretation was developed and added to many machines(Willems *et al* 1987).

Deep learning has diffused throughout the scientific community(Bianchini *et al* 2020). In cardiology, deep learning has been used automated arrythmia classification and has achieved cardiologist level performance(Chen *et al* 2020, Ribeiro *et al* 2020, Nejedly *et al* 2022, Zhao *et al* 2022). The 2020 Physionet/Computing in Cardiology Challenge yielded 212 entries and the most common technique was deep learning and convolutional neural networks(Perez Alday *et al* 2020).

Beyond features immediately identifiable to human interpreters, deep neural networks have been shown to accurately identify hyperkalemia(Galloway *et al* 2019), reduced left ventricular ejection fraction(Attia *et al* 2019b), risk of incident atrial fibrillation(Raghunath *et al* 2021), and age(Attia *et al* 2019a, Lima *et al* 2021, Baek *et al* 2023). Furthermore, an ECG predicted age that exceeds chronological age has been associated with mortality(Lima *et al* 2021, Baek *et al* 2023, Lorenz *et al* 2023, Ladejobi *et al* 2021). However, most models are trained and reported on local data preventing between model comparisons. We sought to compare the performance of models on a new, publicly available data set.

## METHODS

### Data Source, Ethical Declarations, and Reporting Standards

Our data comes from the MIMIC-IV-ECG module(Gow *et al* n.d.) a collection of 800,000 diagnostic electrocardiograms collected at a single hospital and linked to the Medical Information Mart for Intensive Care (MIMIC-IV)(Johnson *et al* n.d., 2023, Baek *et al* 2023) a detailed, deidentified medical record summary of hospitalizations, emergency department visits, and intensive care unit stays. All data is available at PhysioNet(Goldberger *et al* 2000) and was determined to be not human subjects research. This work adheres to the Transparent Reporting of a multivariable prediction model for Individual Prognosis or Diagnosis (TRIPOD) recommendations(Collins *et al* 2015) and the supplementary recommendations from the editors of Respiratory, Sleep, and Critical Care journals(Leisman *et al* 2020).

### Patient Inclusion

MIMIC uses time shifting for patient privacy where the first encounter for each patient is randomly shifted into the future and all dates for that patient are shifted by the same amount. Additionally, patient who are older than 89 at any time during their observation period are further shifted so that no age can be identified, and only relative differences are available. We included all ECGs for all patients with an identifiable (i.e. who were younger than 90 at all their time points) and who were older than 18 at the time of their ECG collection.

### Data Allocation

We allocated the available data by patient with 20% of the patients reserved for final model testing. Of the remaining 80%, we allocated 80% (64% of the total cohort) for model training and 20% (16% of the total cohort) for validation. All ECGs from a subject were included in a single data set. The training set was used for model fitting while the validation set was used for model selection and parameter tuning. The test set was only used for final model evaluation.

### Preprocessing

Individual records were preprocessed by applying a wandering baseline filter (0.2hz wide elliptical infinite impulse response[IIR] high-pass filter centered at 0.8hz with 40db of attenuation), a powerline filter(2^nd^ order digital IIR notch filter centered at 60hz), a 40hz low-pass filter (5^th^ order Butterworth IIR filter), down sampled to 240hz, truncated (or zero padded if required) to 2048 samples, and then standardized (to mean zero and variance 1). Records with missing values were dropped. The signal processing was implemented using the SciPy package(Virtanen *et al* 2020). The 40hz low-pass filter was chosen based on the empiric observation that down sampling produced similar or improved results with a substantial improvement in training times. A comparison of results is shown in eFigure 3.

### Outcomes

Our primary outcomes were age in years at the time ECG acquisition and the patient’s sex recorded in the EHR. In the MIMIC-ECG dataset each subject has a baseline age and an a achor age (The date shifted year in which the baseline age was observed). Each exam has a date shifted timestamp. We calculated the at the age at time of the exam as [Baseline Age] + ([Exam Timestamp Year] - [Anchor Year]).

### Metrics

For the models predicting age our primary metric was mean absolute error (MAE) with additional metrics of mean squared error (MSE) and coefficient of determination (R2). For the models predicting sex we used the maximal Youden’s J-index(Youden 1950) on the validation set to identify the optimal cutoff for classification metrics. The primary evaluation metrics were area under the receiver operating characteristic (AUROC) and accuracy (ACC). Additional metrics were the Brier score(Kruppa *et al* 2014), F1(Buckland and Gey 1994), Sensitivity (Sens)(Altman and Bland 1994), and specificity (Spec)(Altman and Bland 1994). Implementations of all metrics came from the Yardstick package(Kuhn *et al* 2024).

### Models

A schematic representation of all four models is shown in Figure 1. Parameter details, including filter sizes, strides, and max pooling factors, are available in the supplement.

**Figure 1.**
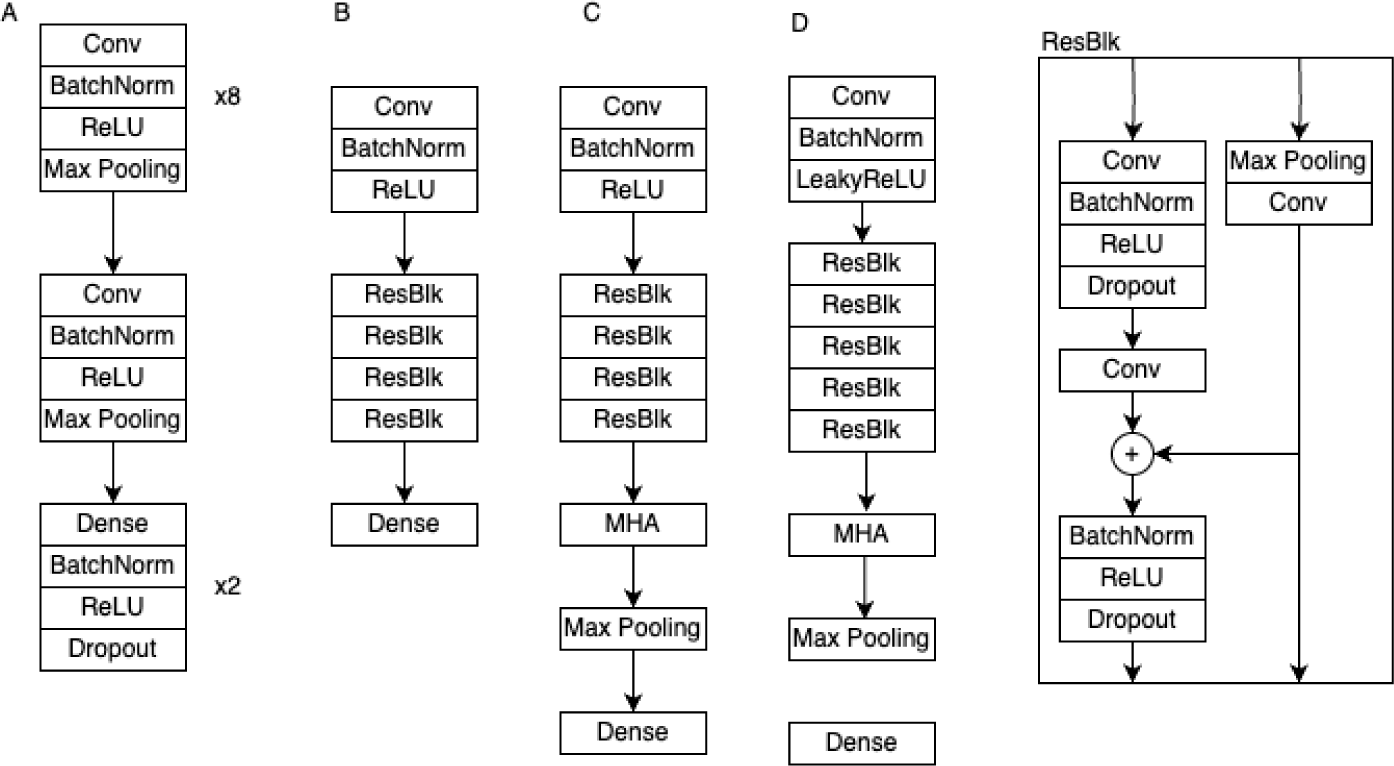
This figure shows the architectures for the a) CNN b) ResNet c) ResNet + MHA and d) MHA models. The ResNet inspired block (ResBlk) is shared by networks B, C, and D.

### Attia (CNN)

Attia and colleagues developed and tested a convolutional neural network (CNN) for the prediction of age and sex(Attia *et al* 2019a) using 774,783 standard ECGs collected at the Mayo Clinic between January 1994 and February 2017. Their model used eight temporal layers, followed by one spatial layer, and then either a classification or regression head. The temporal and spatial layers consisted of 1 dimensional convolutions (either along the time or space axis), batch normalization, ReLU activation, and max pooling. The classification or regression heads consisted of two repeated blocks of fully connected, batch normalization, ReLU activation, and dropout followed by a single output for age or two softmax outputs for sex. The details of kernel sizes, filter counts, strides, and max pooling factors are reported in their original paper(Attia *et al* 2019a) but none of the source code, training data, or model weights are publicly available.

### Lima (ResNet)

Lima and colleagues developed and tested a CNN with residual connections (i.e. ResNet(He *et al* 2016) style) for the prediction of age and associated mortality(Lima *et al* 2021). Their model uses an initial 1-D convolutional block, followed by four ResNet-styled blocks, and finally a dense classification or regression head. The ResNet-styled blocks combine a 1-D convolutional block with a “skip pathway” where input data is added to the convolutional output and then passed forward. The source code, model weights, and most of the training data is publicly available.

### Nejedly (MHA)

Nejedly et. al. submitted the winning entry(Nejedly *et al* 2021) to the 2021 Computing in Cardiology Challenge(Reyna *et al* 2021). They used a CNN with ResNet styled blocks and multi-head attention to deliver the best performance on rhythm classification. We have adapted this model for age and sex prediction. The original source code is publicly available, and we have provided our source code, model weights, and input pipeline for the MIMIC-ECG data.

### Our Model (ResNet+MHA)

Finally, we adapted the Lima model by adding a multi-head attention block after the ResNet styled blocks. We have provided our source code, model weights, and input pipeline for the MIMIC-ECG data.

### Training

All models were trained for 50 epochs with the Adam optimizer and a learning rate of 1e-4 except for the ResNet+MHA model which was trained at 3e-5. The rates were chosen by manual tuning and the observation that the ResNet+MHA model had loss explosion at higher rates.

Each model was trained twice: once for sex prediction (the output predicting the log-odds of male sex) and once for age prediction (with the output predicting age in years). We used binary cross entropy (BCE) as the loss function for sex prediction and mean squared error (MSE) for age prediction. The model with the best validation set results was saved and used for the final evaluation.

### Software and Source Code Availability

We used Python 3.10.12, PyTorch(Paszke *et al* 2019) 2.1.1, and Ignite 0.4.13(Fomin *et al* 2020) on the University of Virginia Rivanna high performance computing cluster to develop and evaluate all models. The source code to support this analysis is available at.

## RESULTS

The final datasets included 510,316 records in the training set, 127,044 records in the validation set, and 162,675 records in the testing set. In the training set 49.1% of the records came from women while 47.8% of the records in the testing set and 49.6% of the records in the validation set came from women. The median age in each of the dataset was 66 and a histogram of observed ages is included in eFigure 1.

All models converged. There was evidence of overfitting in all the sex prediction models with a significant gap between training and validation set performance (Figure 2). In contrast, the gap between training and validation performance was much smaller in the age prediction models.

**Figure 2.**
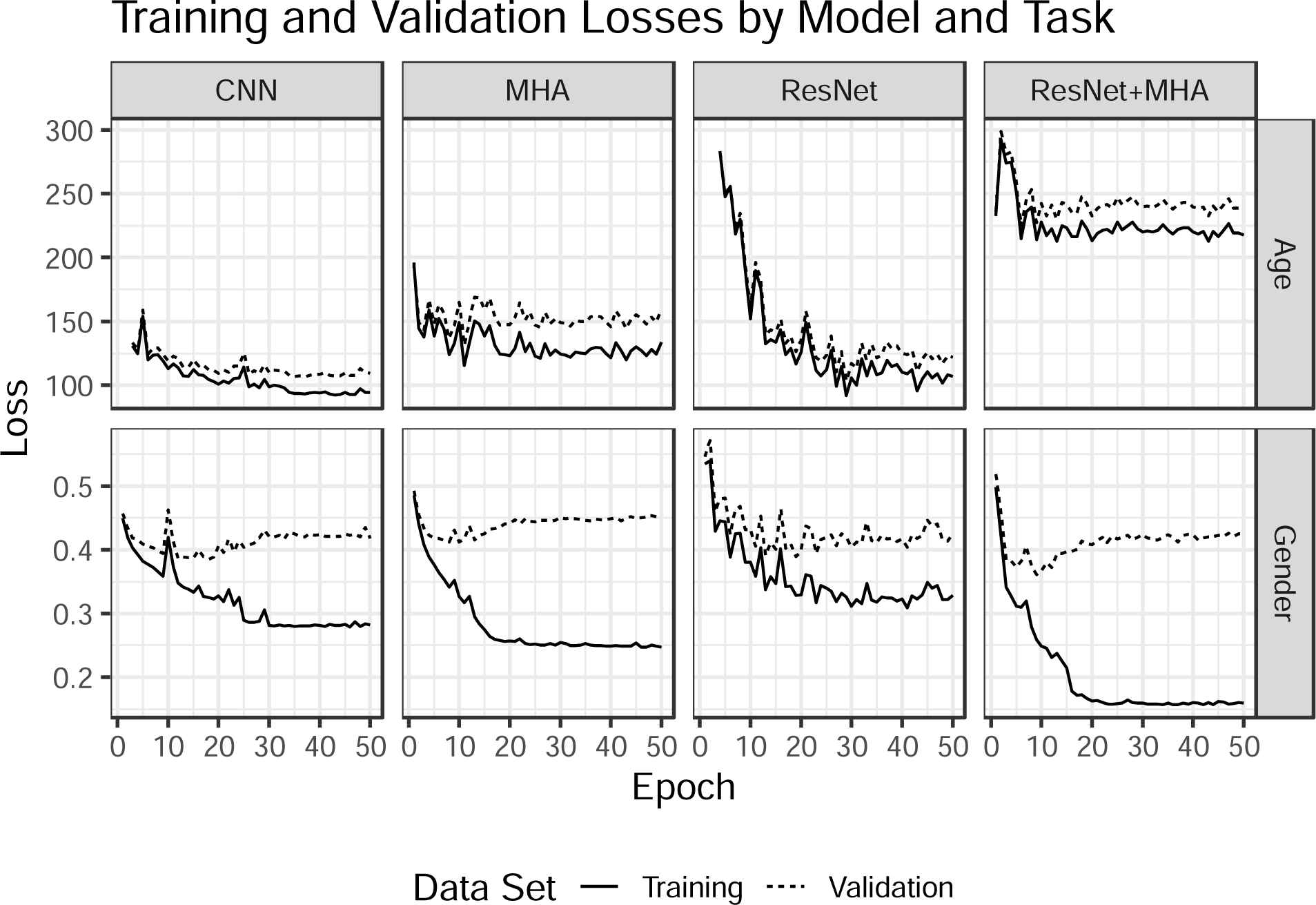
This figure shows the losses by epoch for the age prediction task (top row) and sex prediction task (bottom row). Note the gap between training and validation set performance on the sex prediction task with essentially similar validation set performance across all four models. In contrast, the gap on the age prediction task is generally less than the between model gap.

**Figure 3.**
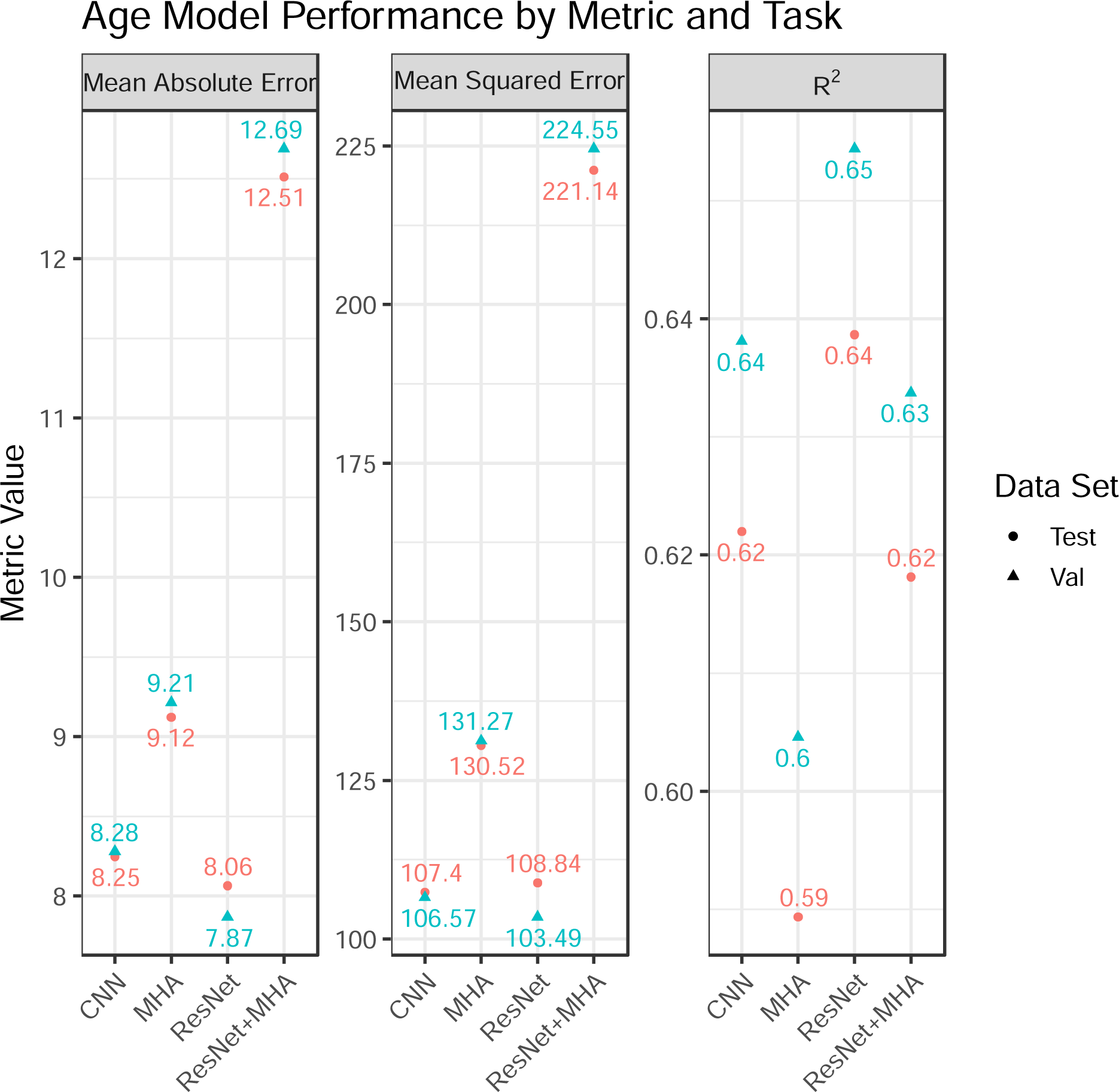
Model performance across the three age prediction metrics. Note that for mean absolute error and mean squared error lower is better while higher is better for R2. The test set is the hold-out set while validation was used for model tuning. Neither set was used for model fitting.

For the task of age prediction, the ResNet model had the best test set MAE and validation set MAE, MSE, and R^2^ while the CNN model had the lowest test set MSE and R^2^(Fgure 3). A plot with observed versus predicted age for a random subset of the data is shown in Figure 4.

**Figure 4.**
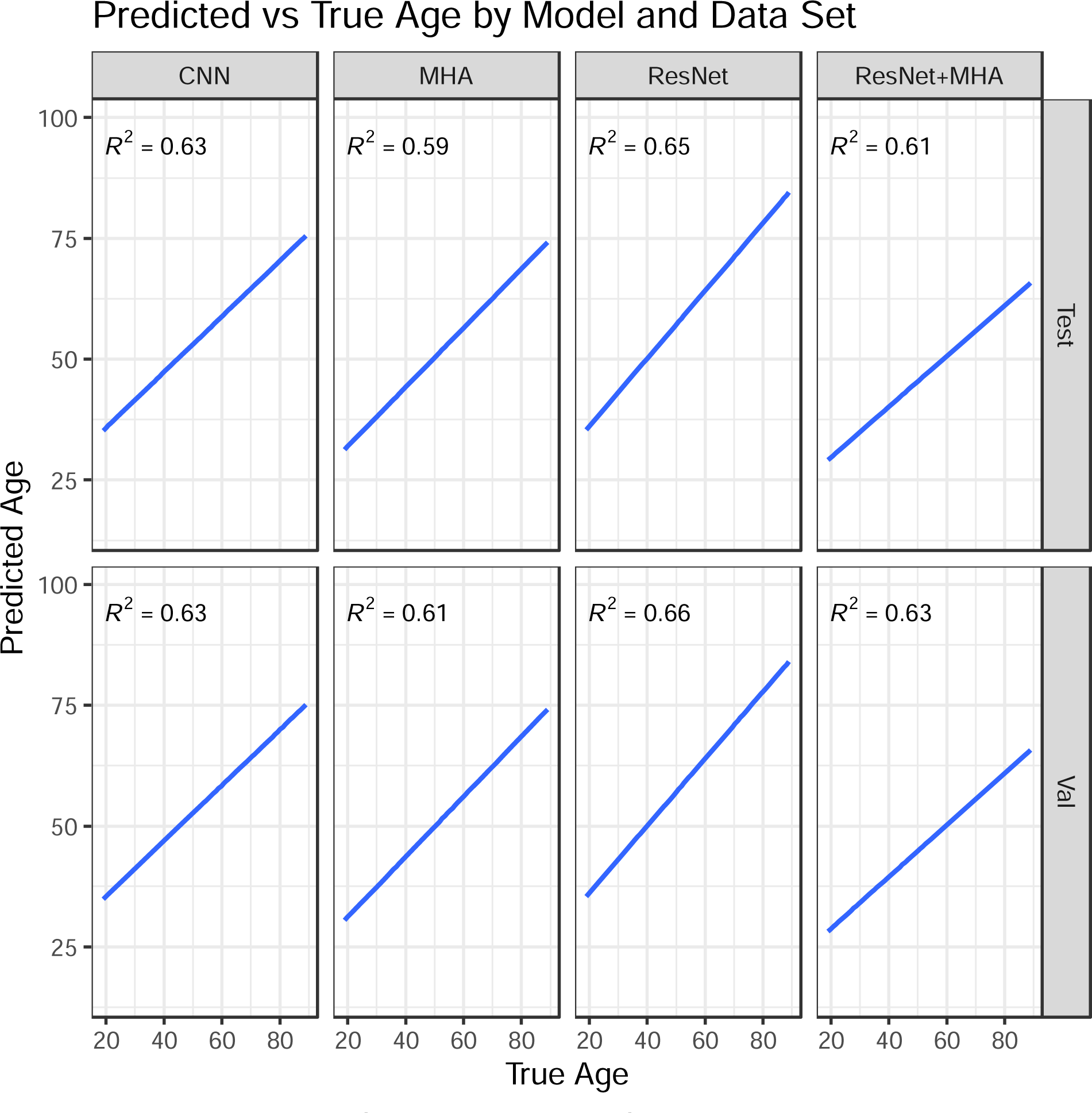
Observed versus predicted ages for a random sample of 10,000 patients across the validation and testing sets. The blue line is the linear regression line of best fit.

**Figure 5.**
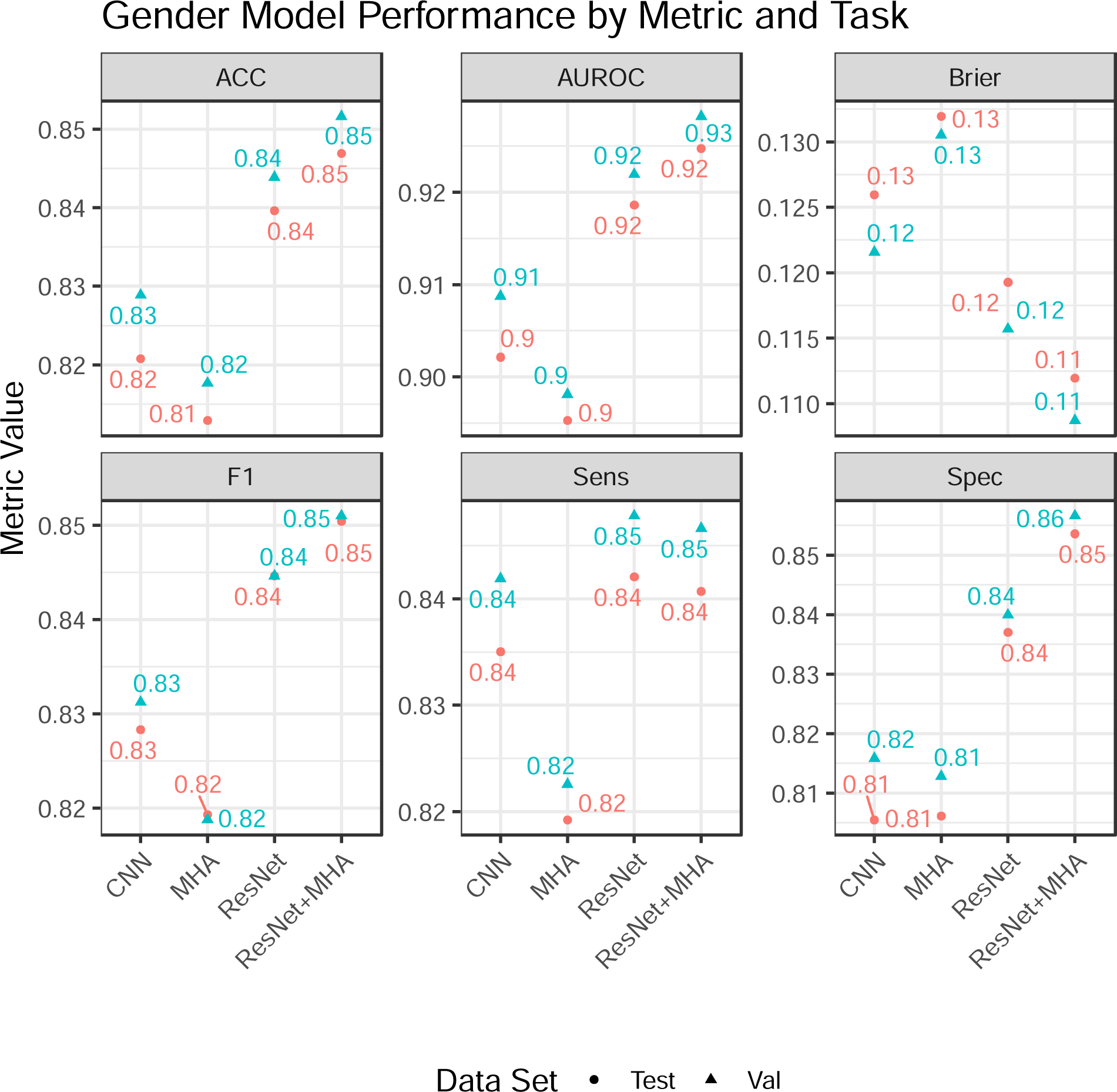
Model performance across the sex prediction metrics: accuracy (ACC), area under the receiver operating curve (AUROC), Brier score, F1, Sensitivity (Sens), and Specificity (Spec). Threshold metrics (ACC, F1, Sens, and Spec) were evaluated at the optimal point selected by Youden’s J- index. The test set is the hold-out set while validation was used for model tuning. Neither set was used for model fitting.

Overall, both models had very similar results. The observed test set performance for the ResNet model was similar to the original published performance (MAE 8.06 years versus 8.38 years) but the CNN performance was worse that published (MAE 8.25 years versus 6.9 years, R^2^ 0.6 versus 0.7). There was visual evidence of heteroskedasticity in the prediction residuals with a decreasing variance for increasing age.

For the task of sex prediction, the ResNet+MHA model had the best accuracy, AUROC, Brier score, F1 score, and specificity on both the testing and validation set (Figure 4). Only the CNN model attempted sex prediction in the prior work. The AUROC for the CNN model was lower than previously published (0.902 versus 0.968) as was the accuracy (0.821 vs 0.904).

## DISCUSSION

We trained and compared four models on a contemporary, freely available dataset. Although age and sex prediction are not directly clinically applicable this framework allows for comparison of architectures across tasks on a single dataset. We acknowledge that only the CNN model was initially designed for sex prediction and that the MHA model was designed for neither sex prediction nor age prediction. However, given that Attia uses the same architecture for both tasks we thought it was reasonable to compare all models on all tasks. An ideal architecture would be foundational and allow for task specific fine-tuning. We found that the best architecture for a sex prediction was not the best for age prediction (and vice versa).

One weakness of our analysis is that we re-implemented parts of all the reference models. The source code for the CNN model was not available and we needed to make some assumptions to develop our implementation. We were able to directly use most of the ResNet implementation but did need to make minor changes due to changes in library versions and data formats. While our implementations are not identical to the original authors, they are a good faith attempt to implement and evaluate the models on a common reference.

A strength of our work includes use of a common open-access dataset and metrics that would allow for direct comparisons of future models against these baselines. This approach is seen in other machine learning domains such as ImageNet(Deng *et al* 2009), SQuAD(Rajpurkar *et al* 2016), and MS-COCO(Lin *et al* 2015) and is partially responsible for the continued progress seen in deep learning.

A significant limitation of both tasks is the lack of direct clinical applicability. Indeed, age prediction in a vacuum is a parlor trick but coupled with real outcomes has potential applications. The observed errors have two parts: the reducible error due to the model and irreducible error due to natural variation. The total amount of reducible error is unknowable and thus we cannot identify when a model is preforming well -- only better than another model. Long term changes in predicted age have been associated with significant outcomes but, to our knowledge, no work has examined if these predictions change over short time scales (say during a hospitalization) and what outcomes these changes might predict. We propose that open, comparable models can facilitate work that examines downstream applications of prediction tasks.

## CONCLUSIONS

Age can be predicted from a standard 12-lead ECG with a performance that varies by network architecture and with significant error. Future models should consider using a publicly available dataset so that between model comparisons can be made.

## DATA AVAILABILITY

The source code for this analysis is available at GitHub. The data is available from PhysioNet.

