## Supplementary Methods for "Age Prediction From 12-lead Electrocardiograms Using Deep Learning: A Comparison of Four Models on a Contemporary, Freely Available Dataset"

### Network Parameters

#### CNN

Temporal Convolutions:

n filters each of 1xk with MaxPool of m

n: 7, 5, 5, 5, 5, 3, 3, 3

k: 16, 16, 32, 32, 64, 64, 64, 64, 64

m: 2, 4, 2, 4, 2, 2, 2, 2

Spatial Convolutions:

128 12x1 filters with

Max Pool of 2

Dense Section:

Layer size: 128, 64

Dropout Rate: 0.8

#### Resnet

n filters of size 17 with resulting sequence length l

n: 128, 196, 256, 320

l: 1024, 256, 64, 16

Residual connection was down sampled through Max Pooling

### MHA

256 filters of size 9 with average pooling (kernel size 2, stride 2)

Multihead attention layer with 8 heads

### Resnet+MHA

n filters of size 17 with resulting sequence length l

n: 128, 196, 256, 320

l: 1024, 256, 64, 16

Residual connection was down sampled through Max Pooling

Multihead attention layer with 4 heads

### Figures

#### eFigure 1

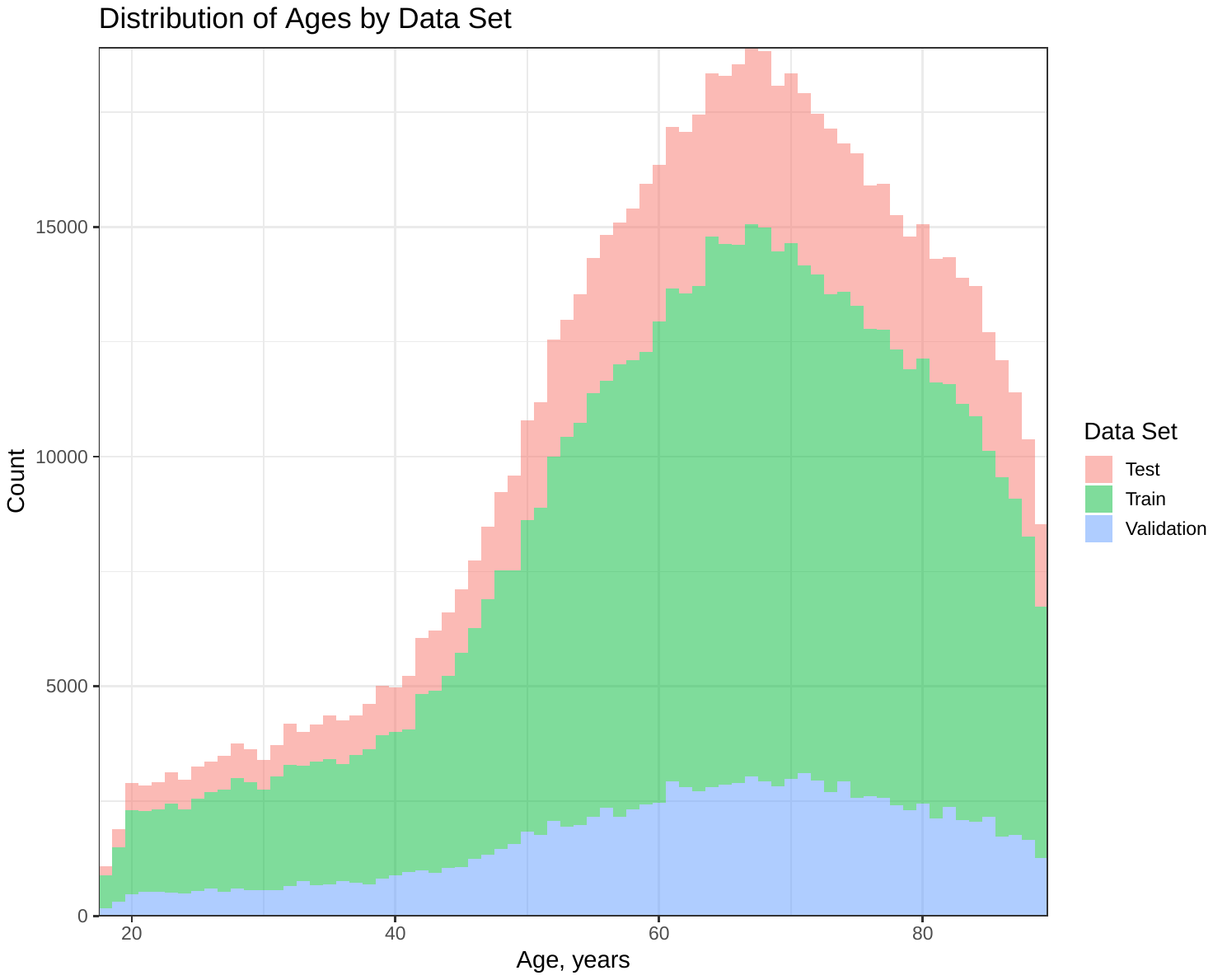

Distribution of exam ages by source data set.

#### eFigure 2

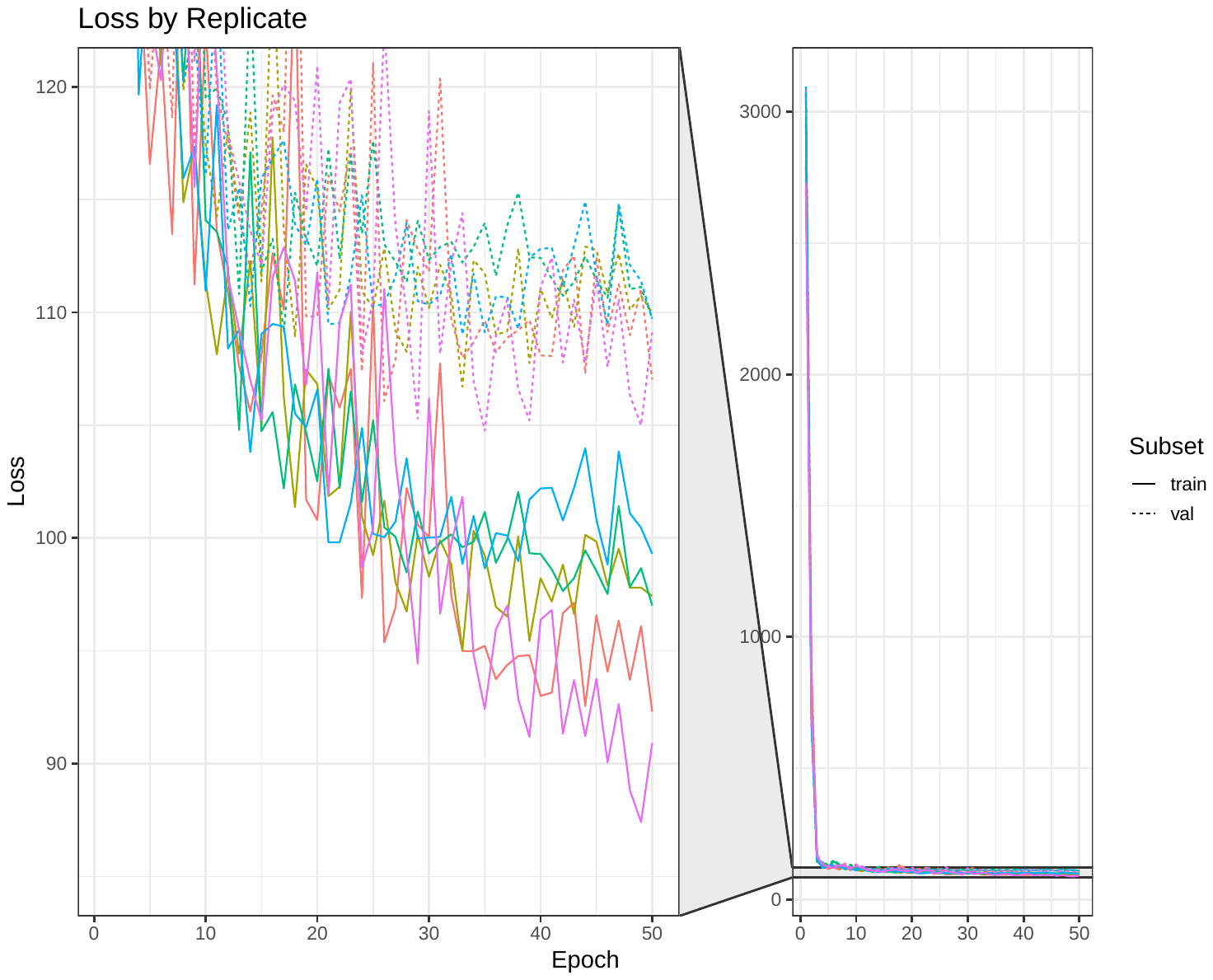

The training (solid line) and validation (dashed line) losses for five replicates of the CNN model.

#### eFigure 3

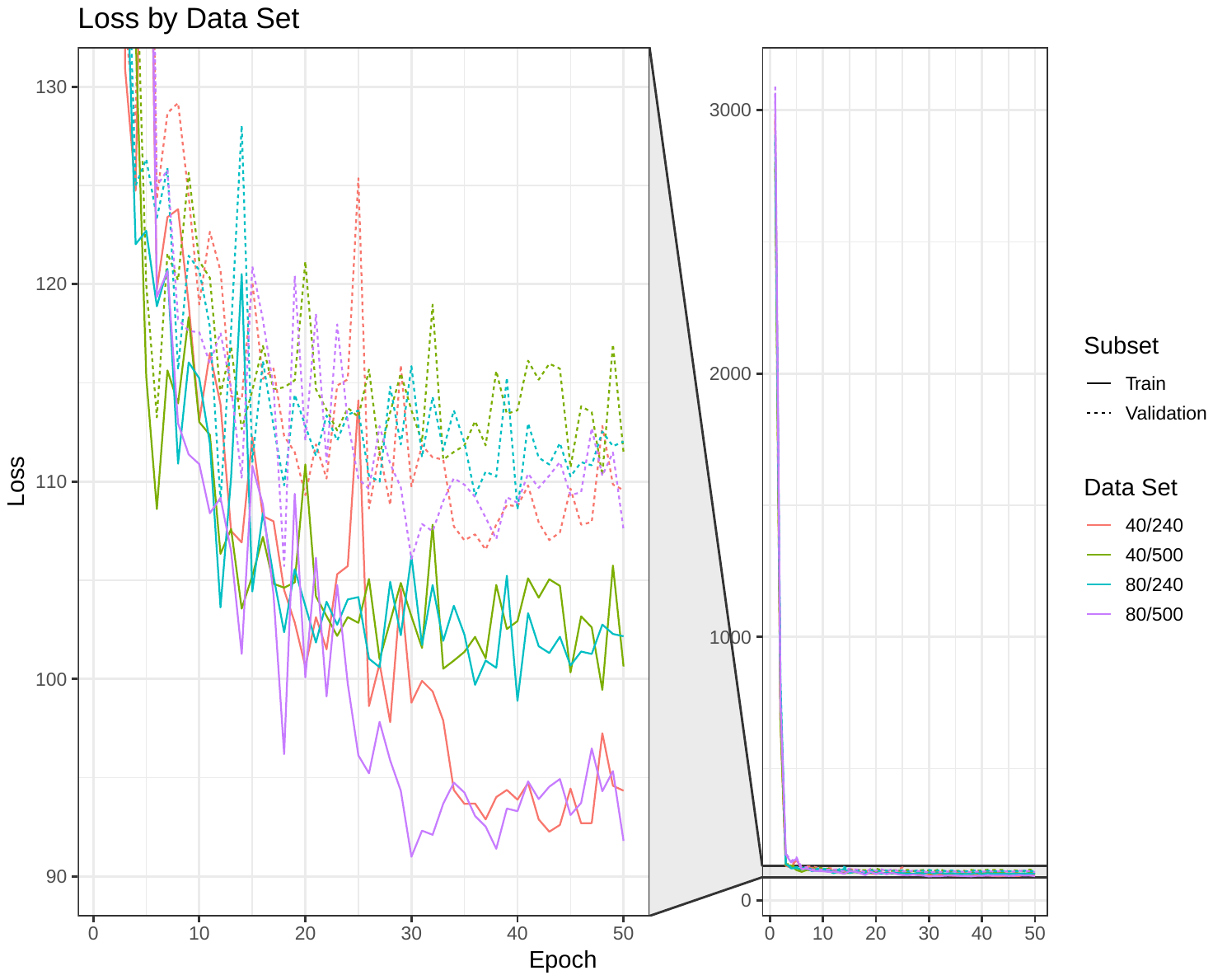

Training and validation losses for the CNN model trained on four data sets: 40/240 (40hz lowpass, downsampled at 240hz), 40/500 (40hz lowpass, source temporal resolution), 80/240 (80hz lowpass, downsampled to 240hz), and 80/500 (80hz lowpass, source temporal resolution). Note that the training loss for 40/240 and 80/500 is similar but the validation loss is modestly improved for 40/240.
